# Risk factors for COVID-19 hospitalization or death during the first Omicron surge in adults: a large population-based case-control study

**DOI:** 10.1101/2022.08.11.22278682

**Authors:** TKT Lo, Hussain Usman, Khokan C. Sikdar, David Strong, Samantha James, Jordan Ross, Lynora M. Saxinger

**Affiliations:** Public Health Surveillance and Informatics, Population and Public Health, Alberta Health Services, Edmonton, Canada; Public Health Evidence and Innovation, Provincial Population and Public Health, Alberta Health Services, Edmonton, Canada; Department of Community Health Sciences, Cumming School of Medicine, University of Calgary, Calgary, Canada; Community Medicine, Population, Public and Indigenous Health, Alberta Health Services, Calgary, Canada; Division of Infectious Diseases, Department of Medicine, University of Alberta, Edmonton, Canada

**Keywords:** Canada, COVID-19, death, hospitalization, intensive-care, male, omicron, vaccine

## Abstract

**Background:** Description of risk factors of severe acute COVID-19 outcomes with the consideration of vaccination status in the era of the Omicron variant of concern are limited.

**Objectives:** To examine the association of age, sex, underlying medical conditions, and COVID-19 vaccination with hospitalization, intensive-care unit (ICU) admission, or death due to the disease, using data from a period when Omicron was the dominant strain.

**Methods:** A population-based case-control study based on administrative health data, that included confirmed COVID-19 patients during January (2022) in Alberta, Canada. Patients who were non-residents, without the provincial healthcare insurance coverage, or ≤18 years of age were excluded. Patients with any severe outcome were the cases; and those without any hospitalization, ICU admission, or death were controls. Adjusted odds ratios, of the explanatory factors of a severe outcome, were estimated using a logistic regression model.

**Results:** There were 90,989 COVID-19 patients included in the analysis; 2% had severe outcomes and 98% were included in the control group. Overall, more COVID patients were found in the younger age-groups (72.0% ≤49 years old), females (56.5%), with no underlying conditions (59.5%), and fully vaccinated patients (90.4%). However, the adjusted odds ratios were highest in the 70–79 age group (28.32; 95% CI 20.6–38.9) or among ≥80 years old (29.8; 21.6–41.0), males (1.4; 1.3–1.6); unvaccinated (16.1; 13.8–18.8), or patients with ≥3 underlying conditions (13.1; 10.9–15.8).

**Conclusion:** Higher risk of severe acute COVID-19 outcomes were associated with older age, the male sex, and increased number of underlying medical conditions. Unvaccination or undervaccination remained as the greatest modifiable risk factor in prevention of severe COVID outcomes. These findings help inform medical decisions and allocation of scarce healthcare resources.

## Background

It is crucial to implement timely and effective public health measures to keep COVID-19 cases and hospitalizations low and to avoid overwhelming the health system. Effective implementation of public health measures depends on the understanding of clinical outcomes of patients infected with SARS-CoV-2, and identification of those who are at the highest risk of severe disease progression. This knowledge can help anticipate clinical resource needs, facilitate preventive intervention distributions, guide therapeutic resource allocations, and support healthcare policymakers’ decisions.

On January 16th, 2022, Health Canada authorized use of the first orally administered antiviral treatment (PAXLOVIDTM; nirmatrelvir/ritonavir) in adult patients with symptomatic COVID-19 to reduce the risk of related severe outcomes (1, 2). Based on the interim results of a phase 2/3 randomized, double-blind study, the new antiviral reduced the risk of COVID-19 related hospitalization or death in unvaccinated high risk adults by 89% compared to placebo (3, 4). The new treatment was estimated to cost USD $530 per course (5), and was expected to be in limited supply relative to the demand (1). A rational allocation, such as to provide access to patients at the highest risk of severe outcomes, is essential to guide effective and transparent distribution of scarce treatments and provide the greatest benefits to Canadians.

A number of risk stratification tools have been developed since the beginning of the COVID-19 pandemic, using data from different populations and settings, to predict various COVID-19 outcomes, including hospitalization, intensive care unit (ICU) admission, and/or death (6-15). These tools range from models that are relatively easy-to use and require fewer, readily available information such as age, sex, and underlying medical conditions (6, 7, 9, 15), to more complex models that require a greater number of parameters including laboratory data and imaging findings (12-14). However, most tools were developed based on data prior to 2021 (6-14) and did not address the impact of vaccination programs and/or new circulating viral variants (16-18). Evidence suggests COVID-19 vaccines significantly lower the risk of SARS-CoV-2 infection, and in particular, confer sustained protection against severe outcome such as hospitalization and mortality (19-21). A model that does not account for vaccination status may overestimate the risk of severe COVID-19 outcomes in those who are fully/partially vaccinated.

Additionally, risk estimation tools developed prior to the emergence of recent variants of concerns (VOCs) including the Delta (B.1.617.2) and Omicron (B.1.1.529) variants would not reflect the current situation. Latest evidence suggests patients infected with the Delta variant had a higher risk of hospitalization compared to the Alpha strain (22), and those infected with Omicron had a lower risk compared to Delta (23-25). Therefore, it is crucial to employ the most up-to-date empirical data for assessment of the contributing factors of severe outcomes, and identification of priority groups for access to scarce resources. These factors may include demographic characteristics, underlying medical conditions, vaccination status, as well as the dominant circulating variant.

## Study objective

The objective of the current study was to examine the factors associated with severe acute COVID-19 outcomes, including hospitalization, intensive care unit (ICU) admission, and/or death using the latest, routinely collected administrative health data.

## Methods

### Design

This study employed a population-based, case-control design. Considered for inclusion were residents in the province of Alberta (Canada) with confirmed COVID-19 diagnosis during the month of January, 2022, when Omicron (BA.1) was the dominant circulating variant. The exclusion criteria were under 18 years of age, prior infection of SARS-CoV-2, and/or hospitalization, ICU admission, or death not due to COVID-19. Diagnosis of COVID-19 was confirmed by a positive result from a transcription-polymerase chain reaction (PCR) test for SARS-CoV-2. Among these patients, those who had any severe outcome (i.e., hospitalization, ICU admission, and/or death) caused by the infection were classified as cases, and those without any severe outcome were put in the control group.

### Data sources

Residents of Alberta have access to publicly funded, universal healthcare services. All Albertans are registered with the Alberta Health Care Insurance Plan except for members of the Armed Forces and the Royal Canadian Mounted Police, federal penitentiary inmates, and residents who have opted out of the insurance plan. Province-wide administrative data captures about 95% of all health system contacts with relatively complete records of the health and healthcare utilization of the Alberta population (26). Alberta Health Services (AHS) is the single health authority for Alberta. Using the data in Communicable Disease and Outbreak Management (CDOM) information system, Provincial Laboratory (ProvLab) database, and other administrative data, AHS conducts COVID-19 surveillance, provides clinical guidelines, and allocate healthcare resources. We accessed these data to identify the cases and non-cases.

The Immunization and Adverse Reactions to Immunization (Meditech/ ImmARI) data were used to determine the COVID-19 vaccination status of individuals. Each person registered with the provincial insurance plan has a 9-digit unique lifetime identifier (ULI), we used this key for deterministic linkage of all data at the patient level. The underlying health conditions of the patients were determined from the diagnoses contained in administrative data including the Alberta Physician Billing Claims, National Ambulatory Care Reporting System (NACRS), and Discharge Abstract Database (DAD). The claims have information about physician visits and diagnoses during the service episodes, i.e., coded in the 9^th^ Revision of Clinical Modification of the International Classification of Diseases (ICD-9-CM). The NACRS and the DAD contain ambulatory visits and hospital admissions information, respectively. Diagnoses during each visit (or hospital stay) were coded using the enhanced Canadian version of the 10^th^ revision of ICD (ICD-10-CA). Finally, information of death and underlying cause (coded in ICD-10) was extracted from the vital statistics. These data sources have been used in other health and health services research (27-30). Validity of information in these data sources were reported elsewhere (26, 31).

### Outcome measure

Derived from CDOM data, the outcome measure was any COVID-19 related hospitalization, ICU admission, and/or death indicating a progression of severe acute illness.

### Other variables

Inclusion of covariates in our risk estimation model was guided by other published reports (i.e., variables identified in the literature as predictors of COVID-19 outcomes) (32-37), as well as the accessibility of data when the model is applied by practitioners in the community setting (e.g., family physicians). Accordingly, our team selected age, sex, underlying medical conditions, and vaccination status for the analysis.

For underlying medical conditions, we included 24 comorbidities that have been associated with a higher risk of SARS-CoV-2 infection and/or severe health outcomes (38). They were: a. respiratory diseases (asthma, COPD, other chronic lung disease); b. heart diseases (heart failure, ischemic heart disease, hypertension, other heart disease); c. cerebrovascular diseases (stroke, other cerebrovascular disease); d. metabolic diseases (diabetes type I, diabetes type II, diabetes due to underlying condition or other specified diabetes, and other metabolic disease); and e. other (clinical obesity, clinically underweight, renal disease, liver disease, blood disorder, other immunosuppression, organ transplant, cancer, dementia & Alzheimer’s disease, neurological/musculoskeletal disorder, and Down’s syndrome). These high risk conditions conform to those of a published COVID-19 vaccine effectiveness study by Thompson et al. (38), who also published condition identification algorithms (39). Briefly, identification of the each medical condition were based on the diagnoses coded in International Classification of Diseases 9th revision (ICD-9) and 10th revision (ICD-10). A person with at least one physician visit, ambulatory visit, or hospital admission with the applicable ICD diagnosis code recorded in the previous 24 months was considered to have that medical condition. We used the approach of Knight et al. and applied a count of the high risk conditions in our analysis (13). The number of medical conditions have shown to be significantly associated with COVID-19 outcomes (10, 13, 40).

Vaccination status was classified into five categories: a) unvaccinated, b) one-dose, c) two-dose and >6 months from the second dose, d) two-dose and ≤6 months from the second dose, and e) three or more doses. We created two categories for patients with two doses because evidence suggests that protection from COVID-19 vaccines may wane (i.e., even after two doses or being fully vaccinated) after six months (41).

### Statistical analysis

Demographic characteristics of the cases and controls were summarized using means and standard deviations (continuous variables) and proportions (categorical variables). Examination of factors associated with severe outcomes was based on multivariable logistic regression modeling. In the model, the dependent variable denoted any severe outcome (i.e., hospitalization, ICU admission, and/or death); the independent variables were age, sex, underlying conditions, and vaccination status. The probabilities of severe outcome were estimated using the formula: *p*= *e*^(*a*+*b*)/(1+*e*^(*a*+*b*)), where “*a*”represents the intercept and “*b*”represents the coefficients of the regression models. All statistical analyses were performed in SAS 9.04 (SAS Institute Inc. NC, USA). Statistical significance was defined as P < 0.05.

### Ethics

Ethics approvals were obtained from the Conjoint Health Research Ethics Board of Alberta (REB22-0242) who waived the requirement of informed consent.

## Results

In Alberta, 118,456 persons had confirmed SARS-CoV-2 infection between the first and 31^st^ of January, 2022. We included 90,989 (76.8%) COVID-19 patients in our analytic sample. The flow of the inclusion and exclusion of patients is illustrated in **Figure 1**.

**Figure 1:**
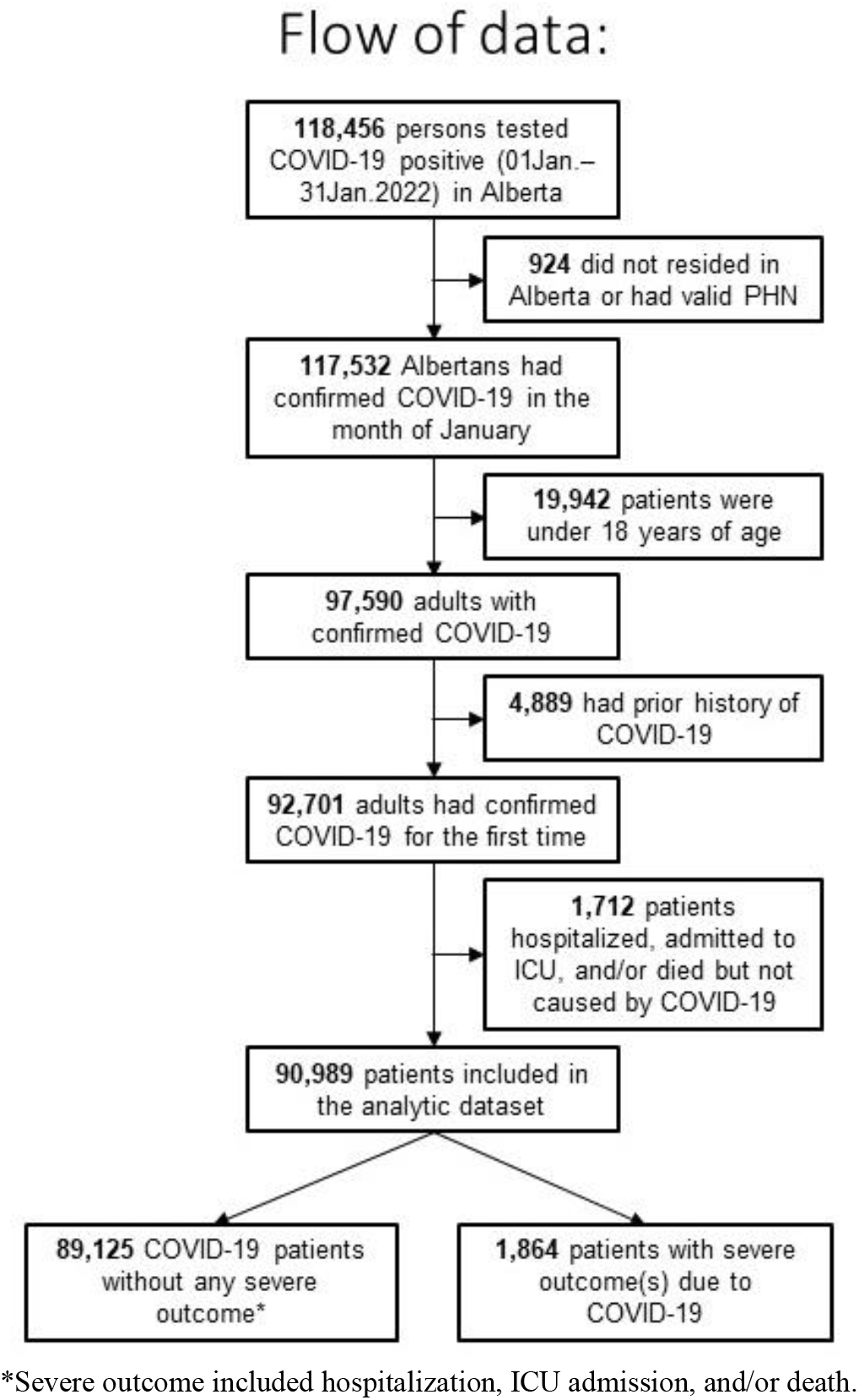
Flow chart of COVID-19 patients with and with severe outcome

Among these COVID-19 patients, SARS-CoV-2 virus typing was available from 39.9% of the cases; 99.1% (35,970 of 36,306) were the Omicron (BA.1) variant and 0.9% was the Delta variant. There were 1,718 patients hospitalized (1.9%), 246 (0.3%) admitted to the ICU, and 382 (0.4%) died. A total of 1,864 patients (2.0%) had one or more severe acute outcomes due to the disease, and 89,125 patients (98.0%) COVID-19 patients had no hospitalization or ICU admission, or not died.

More patients in our analytic sample were female (56.5%); the mean age (standard deviation) was 42.04 (16.43) years. More COVID-19 patients (72.0%) were in the younger age groups (18–49 years). A total of 204,517 vaccination records were linked to our sample. The majority of doses (97.3%) were mRNA vaccines (i.e., Pfizer-BioNTech or Moderna mRNA-1273) and 2.2% were Oxford-AstraZeneca (Vaxzevria). Over 90% of confirmed COVID-19 patients were fully vaccinated (i.e., ≥2 doses).

The top five underlying conditions in the sample were hypertension, other metabolic disease, asthma, clinical obesity, and ischemic heart disease. However, most patients (59.5%) did not have any high risk medical condition. Characteristics of the sample are describe in more detail in **Table 1**.

**Table 1:** Characteristics of patients with a confirmed COVID-19 diagnosis during January (2022) in Alberta, Canada

| Characteristics | Number | Proportion (%) |
| --- | --- | --- |
| Age group |  |  |
| 18-29 | 22,676 | 24.92 |
| 30-39 | 22,827 | 25.09 |
| 40-49 | 20,031 | 22.01 |
| 50-59 | 12,908 | 14.19 |
| 60-69 | 6,315 | 6.94 |
| 70-79 | 2,740 | 3.01 |
| 80&up | 3,492 | 3.84 |
| Sex |  |  |
| Female | 51,435 | 56.53 |
| Male | 39,554 | 43.47 |
| Vaccination status |  |  |
| Unvaccinated | 7,047 | 7.74 |
| 1-dose | 1,665 | 1.83 |
| 2-dose, ≥6mo | 37,847 | 41.60 |
| 2-dose, <6mo | 24,760 | 27.21 |
| 3-dose | 19,670 | 21.62 |
| No. underlying conditions<sup>†</sup> |  |  |
| 0 | 54,102 | 59.46 |
| 1 | 19,958 | 21.93 |
| 2 | 8,669 | 9.53 |
| 3+ | 8,260 | 9.08 |
| Any severe outcome* |  |  |
| No | 89,125 | 97.95 |
| Yes | 1,864 | 2.05 |
| Total patients included | 90,989 | 100.00 |

\*Severe outcomes include hospitalization, ICU admission, and/or death due to COVID-19. Conditions were asthma, COPD, other chronic lung disease, heart failure, ischemic heart disease, hypertension, other heart disease, stroke, other cerebrovascular disease, diabetes I, diabetes II, diabetes due to underlying condition, other metabolic disease, clinical obesity, clinically underweight, renal disease, liver disease, blood disorder, other immunosuppression, organ transplant, cancer, dementia & Alzheimer's disease, neurological/ musculoskeletal disorder, and Down's syndrome.

In logistic regression analysis, chi-squared test against the null hypothesis (i.e., at least one of the explanatory variable in the model was not equal to zero) was significant (P <0.001). Each of the included explanatory variables were also statistically significant; the adjusted odds ratios and chi-squared statistics are listed in **Table 2** and illustrated in **Figure 2**. Model’s R-squared statistic was estimated at 37.85%. Based on model intercept and coefficients, the probabilities of severe outcome were calculated. The probability of a COVID-19 severe outcome was higher in older age groups, males, undervaccinated (≤1 dose), and/or patients with more underlying medical conditions; see **Table 3** for details.

**Table 2:** Results of multivariable logistic regression on severe outcome^†^ due to COVID-19

| Variable | Odds ratio (95% CI) | Coeff. | Wald ChiSq | P |  |
| --- | --- | --- | --- | --- | --- |
| Intercept | NA | -7.8692 | 2366.10 | <.0001 | * |
| Age group |  |  |  |  |  |
| 18-29 | Reference |  |  |  |  |
| 30-39 | 1.40 (0.99–1.99) | 0.3377 | 3.55 | 0.0596 |  |
| 40-49 | 2.07 (1.49–2.88) | 0.7268 | 18.50 | <.0001 | * |
| 50-59 | 4.07 (2.96–5.59) | 1.4037 | 74.94 | <.0001 | * |
| 60-69 | 9.59 (7.01–13.12) | 2.2606 | 199.34 | <.0001 | * |
| 70-79 | 28.32 (20.63–38.89) | 3.3438 | 427.36 | <.0001 | * |
| 80&up | 29.77 (21.63–40.98) | 3.3936 | 433.10 | <.0001 | * |
| Sex |  |  |  |  |  |
| Female | Reference |  |  |  |  |
| Male | 1.44 (1.30–1.60) | 0.3664 | 47.76 | <.0001 | * |
| Vaccination status |  |  |  |  |  |
| 3-dose | Reference |  |  |  |  |
| Unvaccinated | 16.12 (13.81–18.82) | 2.7803 | 1238.66 | <.0001 | * |
| 1-dose | 6.21 (4.62–8.36) | 1.8267 | 145.22 | <.0001 | * |
| 2-dose, ≥6mo | 2.26 (1.96–2.60) | 0.8138 | 125.28 | <.0001 | * |
| 2-dose, <6mo | 2.35 (1.96–2.82) | 0.8560 | 85.54 | <.0001 | * |
| Underlying conditions<sup>‡</sup> |  |  |  |  |  |
| 0 | Reference |  |  |  |  |
| 1 | 2.62 (2.14–3.19) | 0.9618 | 89.72 | <.0001 | * |
| 2 | 4.80 (3.92–5.88) | 1.5683 | 228.64 | <.0001 | * |
| 3+ | 13.12 (10.90–15.81) | 2.5745 | 736.79 | <.0001 | * |

\*Statistically significant. <sup>†</sup>Severe outcomes include hospitalization, ICU admission, and/or death due to COVID-19. <sup>‡</sup>Conditions were asthma, COPD, other chronic lung disease, heart failure, ischemic heart disease, hypertension, other heart disease, stroke, other cerebrovascular disease, diabetes I, diabetes II, diabetes due to underlying condition, other metabolic disease, clinical obesity, clinically underweight, renal disease, liver disease, blood disorder, other immunosuppression, organ transplant, cancer, dementia & Alzheimer's disease, neurological/ musculoskeletal disorder, and Down's syndrome. Abbreviations: Coeff=coefficient, ChiSq=Chi-squared statistic, CI=confidence interval

**Table 3:**
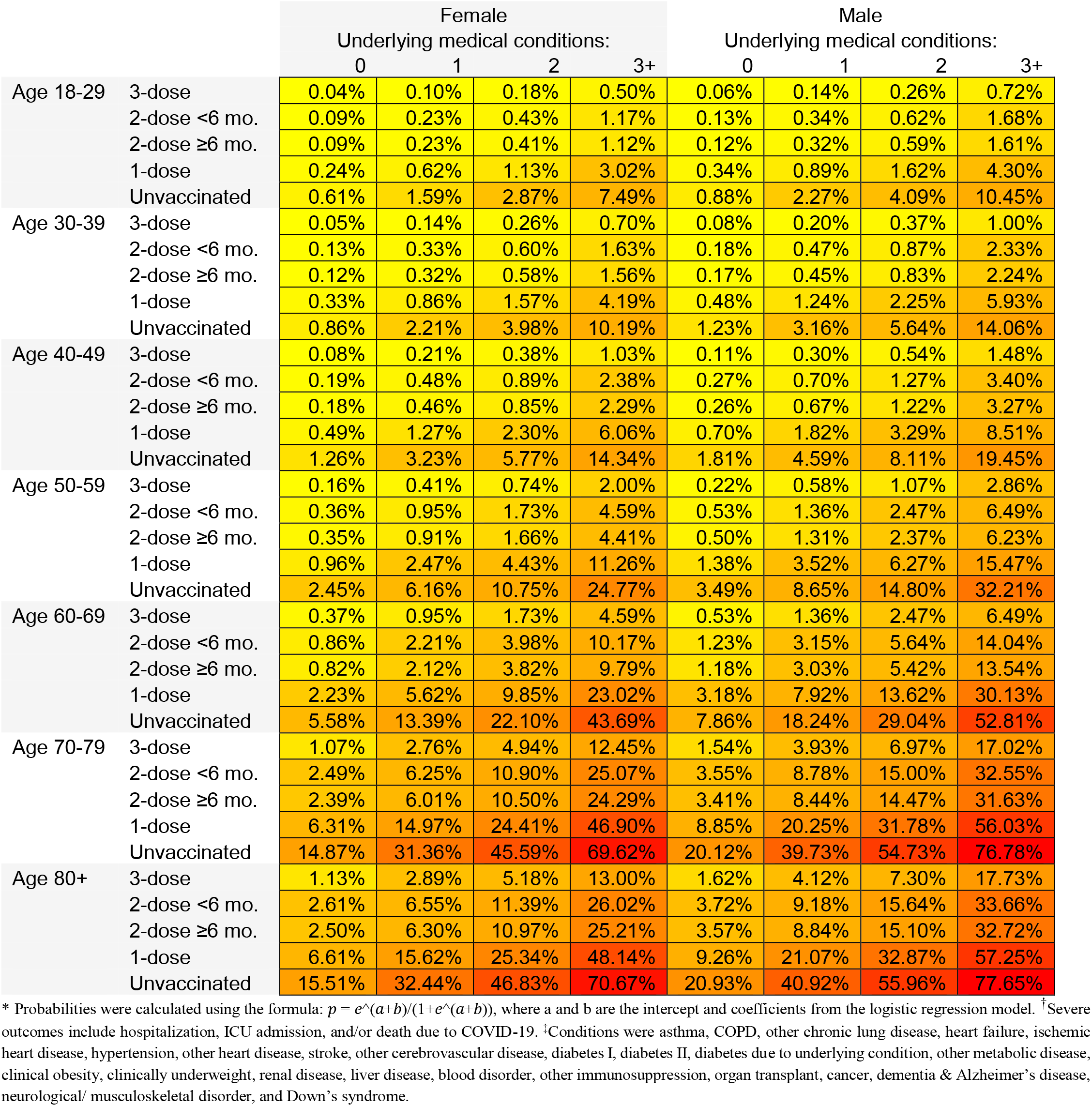
Estimated probability* of severe outcome^†^ due to COVID-19 by age-group, vaccination status, and number of conditions

|  |  | Female |  |  |  | Male |  |  |  |
| --- | --- | --- | --- | --- | --- | --- | --- | --- | --- |
|  |  | Underlying medical conditions: |  |  |  | Underlying medical conditions: |  |  |  |
|  |  | 0 | 1 | 2 | 3+ | 0 | 1 | 2 | 3+ |
| Age 18-29 | 3-dose | 0.04% | 0.10% | 0.18% | 0.50% | 0.06% | 0.14% | 0.26% | 0.72% |
|  | 2-dose <6 mo. | 0.09% | 0.23% | 0.43% | 1.17% | 0.13% | 0.34% | 0.62% | 1.68% |
|  | 2-dose ≥6 mo. | 0.09% | 0.23% | 0.41% | 1.12% | 0.12% | 0.32% | 0.59% | 1.61% |
|  | 1-dose | 0.24% | 0.62% | 1.13% | 3.02% | 0.34% | 0.89% | 1.62% | 4.30% |
|  | Unvaccinated | 0.61% | 1.59% | 2.87% | 7.49% | 0.88% | 2.27% | 4.09% | 10.45% |
| Age 30-39 | 3-dose | 0.05% | 0.14% | 0.26% | 0.70% | 0.08% | 0.20% | 0.37% | 1.00% |
|  | 2-dose <6 mo. | 0.13% | 0.33% | 0.60% | 1.63% | 0.18% | 0.47% | 0.87% | 2.33% |
|  | 2-dose ≥6 mo. | 0.12% | 0.32% | 0.58% | 1.56% | 0.17% | 0.45% | 0.83% | 2.24% |
|  | 1-dose | 0.33% | 0.86% | 1.57% | 4.19% | 0.48% | 1.24% | 2.25% | 5.93% |
|  | Unvaccinated | 0.86% | 2.21% | 3.98% | 10.19% | 1.23% | 3.16% | 5.64% | 14.06% |
| Age 40-49 | 3-dose | 0.08% | 0.21% | 0.38% | 1.03% | 0.11% | 0.30% | 0.54% | 1.48% |
|  | 2-dose <6 mo. | 0.19% | 0.48% | 0.89% | 2.38% | 0.27% | 0.70% | 1.27% | 3.40% |
|  | 2-dose ≥6 mo. | 0.18% | 0.46% | 0.85% | 2.29% | 0.26% | 0.67% | 1.22% | 3.27% |
|  | 1-dose | 0.49% | 1.27% | 2.30% | 6.06% | 0.70% | 1.82% | 3.29% | 8.51% |
|  | Unvaccinated | 1.26% | 3.23% | 5.77% | 14.34% | 1.81% | 4.59% | 8.11% | 19.45% |
| Age 50-59 | 3-dose | 0.16% | 0.41% | 0.74% | 2.00% | 0.22% | 0.58% | 1.07% | 2.86% |
|  | 2-dose <6 mo. | 0.36% | 0.95% | 1.73% | 4.59% | 0.53% | 1.36% | 2.47% | 6.49% |
|  | 2-dose ≥6 mo. | 0.35% | 0.91% | 1.66% | 4.41% | 0.50% | 1.31% | 2.37% | 6.23% |
|  | 1-dose | 0.96% | 2.47% | 4.43% | 11.26% | 1.38% | 3.52% | 6.27% | 15.47% |
|  | Unvaccinated | 2.45% | 6.16% | 10.75% | 24.77% | 3.49% | 8.65% | 14.80% | 32.21% |
| Age 60-69 | 3-dose | 0.37% | 0.95% | 1.73% | 4.59% | 0.53% | 1.36% | 2.47% | 6.49% |
|  | 2-dose <6 mo. | 0.86% | 2.21% | 3.98% | 10.17% | 1.23% | 3.15% | 5.64% | 14.04% |
|  | 2-dose ≥6 mo. | 0.82% | 2.12% | 3.82% | 9.79% | 1.18% | 3.03% | 5.42% | 13.54% |
|  | 1-dose | 2.23% | 5.62% | 9.85% | 23.02% | 3.18% | 7.92% | 13.62% | 30.13% |
|  | Unvaccinated | 5.58% | 13.39% | 22.10% | 43.69% | 7.86% | 18.24% | 29.04% | 52.81% |
| Age 70-79 | 3-dose | 1.07% | 2.76% | 4.94% | 12.45% | 1.54% | 3.93% | 6.97% | 17.02% |
|  | 2-dose <6 mo. | 2.49% | 6.25% | 10.90% | 25.07% | 3.55% | 8.78% | 15.00% | 32.55% |
|  | 2-dose ≥6 mo. | 2.39% | 6.01% | 10.50% | 24.29% | 3.41% | 8.44% | 14.47% | 31.63% |
|  | 1-dose | 6.31% | 14.97% | 24.41% | 46.90% | 8.85% | 20.25% | 31.78% | 56.03% |
|  | Unvaccinated | 14.87% | 31.36% | 45.59% | 69.62% | 20.12% | 39.73% | 54.73% | 76.78% |
| Age 80+ | 3-dose | 1.13% | 2.89% | 5.18% | 13.00% | 1.62% | 4.12% | 7.30% | 17.73% |
|  | 2-dose <6 mo. | 2.61% | 6.55% | 11.39% | 26.02% | 3.72% | 9.18% | 15.64% | 33.66% |
|  | 2-dose ≥6 mo. | 2.50% | 6.30% | 10.97% | 25.21% | 3.57% | 8.84% | 15.10% | 32.72% |
|  | 1-dose | 6.61% | 15.62% | 25.34% | 48.14% | 9.26% | 21.07% | 32.87% | 57.25% |
|  | Unvaccinated | 15.51% | 32.44% | 46.83% | 70.67% | 20.93% | 40.92% | 55.96% | 77.65% |

\* Probabilities were calculated using the formula: $p = e^{(a+b)} / (1 + e^{(a+b)})$ , where a and b are the intercept and coefficients from the logistic regression model. † Severe outcomes include hospitalization, ICU admission, and/or death due to COVID-19. ‡ Conditions were asthma, COPD, other chronic lung disease, heart failure, ischemic heart disease, hypertension, other heart disease, stroke, other cerebrovascular disease, diabetes I, diabetes II, diabetes due to underlying condition, other metabolic disease, clinical obesity, clinically underweight, renal disease, liver disease, blood disorder, other immunosuppression, organ transplant, cancer, dementia & Alzheimer's disease, neurological/ musculoskeletal disorder, and Down's syndrome.

**Figure 2:**
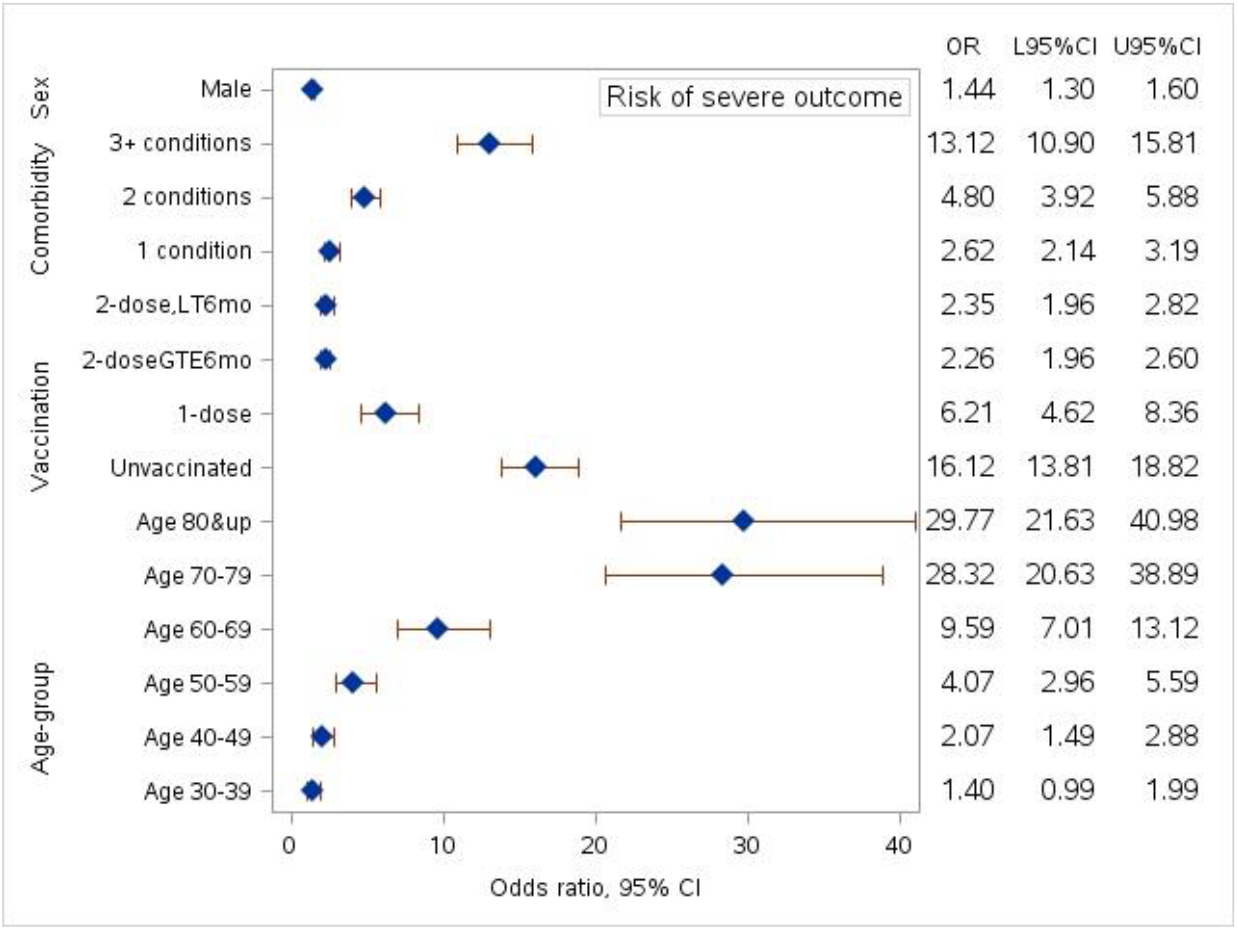
Odds ratios from multivariable logistic regression analysis

## Discussion

### Principal findings

We quantified the risk of severe outcome (i.e., hospitalization, ICU admission, and/or death) due to COVID-19 using age, sex, number of underlying conditions, and vaccination status of the patients. Despite more confirmed cases were found among the younger age, fully vaccinated, or with no comorbidity groups during the early part of the Omicron wave, the adjusted risks were notably higher in patients who were older, not vaccinated or had only 1-dose, and/or with underlying medical conditions. Specifically, COVID-19 patients who were ≥70 years of age had over 28.3x greater risk of severe outcome compared to those 18–29 years of age, patients who had ≥3 high risk medical conditions had 13.1x higher risk than those without any underlying conditions, and unvaccinated patients had an 16.1x increased risk compared to those who had their booster shots (i.e., >2 doses). We also found that males had a 1.4x higher risk than females. Based on these results, a person between 25 years of age, with no underlying medical condition, three doses of vaccine, and female would have about 0% chance of severe acute COVID-19 outcomes. Whereas another person at 70 years old, with 3 comorbid conditions, unvaccinated, and male would have a marked 77% chance of severe outcome.

These findings provide insights into some notable risk factors of COVID-19 severe outcomes. Most importantly, our results indicate that vaccination continues to be a significant and modifiable factor of hospitalization, ICU admission, or death due to COVID-19. Because over 97% of COVID-19 vaccines administered in Alberta were mRNA vaccines, and patients with only two doses had over 2x greater risk of severe outcome than 3-doses (i.e., a booster shot), our results also suggest possible waning of COVID-19 vaccine protection over time (5). Nevertheless, the estimated ORs and 95% CIs for the two categories of 2-doses (i.e., <6 months or ≥6 months from the last dose) did not suggest significantly differences at the 6-month cut-off. Future research should further investigate the best timing for a booster dose.

### Comparison with other studies

Comparisons of our results with those previously reported should be made with caution due to the differences in population characteristics (e.g., ethnic/race composition), circulating SARS-CoV-2 virus strains, explanatory variables (e.g., with or without individuals’ vaccination status), and the outcome measure. Nevertheless, our results seems to be in agreement with earlier studies. Yamada et al. found that COVID-19 patients who were 75 years or older had a 2.0x greater risk of respiratory failure than those who were younger (11). Vila-Corcoles et al. shown that COVID-19 patients who were 80 years or older had 10.9 times the risk of ICU admission or death compared to those in the 50–64 years age group (9). Furthermore the results from Diep et al. suggest that with each year increase in age, the risk of hospitalization due to COVID-19 increases 1.1x (6). However, our results indicate that a COVID-19 severe outcome might not have a linear relationship with age (see illustration in **Figure 2**); further studies into their relationship are warranted.

Other studies have found that the risk of a COVID-19 severe outcome increased with the number of underlying medical conditions and the male sex (10, 13, 40). Particularly, a large US cohort study found that males have a 1.5x greater risk of COVID-19 hospitalization than females (10). Two other European studies show that males have a 1.7x and 3.4x increased hospitalization risk in the Spanish and Belgian populations, respectively (6, 9). In a Japanese population-based study, males were also found to have a 1.7x higher risk of hospitalization due to COVID-19 then females (11). After adjusting for age, medical conditions, and vaccination status, our results (i.e., a 1.4x increased risk for the males) seems to be consistent with these findings.

### Strength & Limitations

There are several strengths and limitations of this study. This study was based on a population-based design and included a large sample; these attributes contribute to the internal validity of the results. The number of variables included in the model also represents a strength. Our model does not involve laboratory or radiological data, but only requires information that is more readily available to the medical practitioners. Hence, this model represents a simple, risk-stratifying tool that can be easy-to-use in the community at the point of test positivity.

The novelty of this study also lies in the inclusion of patients’ vaccination status in our risk calculation and employed up-to-date data when Omicron was (and still is at the time of writing this report) the dominant variant. Because COVID-19 vaccines significantly protect patients from severe outcome even after a SARS-CoV-2 infection, and patients’ outcome varies by the virus strains, inclusion of vaccination status and use of the latest empirical data contributes to a more accurate estimation of the current risks. However, using data from an Omicron dominant period may also represent a limitation. Results that are based on these data may not be generalizable to other periods when Omicron (BA.1) is not the dominant strain. As the SARS-CoV-2 virus evolves over time, continuous study of the risk of severe COVID-19 outcomes is essential (42).

Lastly, the inclusion of confirmed COVID-19 cases may have introduced some selection bias. As of January, 2020, Alberta PCR testing became less accessible to the general public due to a sharp increase in demand, and was only accessible to eligible individuals with higher risk of SARS-CoV-2 infection and related outcomes (43). These individuals included healthcare workers, household member of a healthcare worker, persons living in long-term care, older age and/or indigenous, those with certain health conditions (e.g., diabetes, obesity, chronic kidney disease, congestive heart failure, COPD, asthma, and immunocompromised), and international travelers. Consequently, our sample might have included more individuals who were at a higher risk of infection while others infected in the community with mild (or no) symptoms but not tested could be missed. Although our risk calculation was adjusted to several factors, including high risk medical conditions, residual confounding is possible. Interpretation of our results must be made within the context of these limitations.

### Implication of findings

Our results in the probability table. i.e., **Table 3**, can be used by practitioners as a pragmatic tool to identify patients with the highest risk of severe outcome at the time of COVID-19 diagnosis and guide medical decision-makings. Based on the findings, therapeutic interventions and other scarce resources can be effectively distributed to patients who are likely to benefit the most. The results from our model can also be programmed into an online risk-calculator for convenient and quick access of these information.

## Conclusions

This study examined age, sex, underlying medical conditions, and vaccination status and their association with severe COVID-19 outcomes including hospitalization, ICU admission, and death. A severe acute outcome was associated with older age, male sex, and more underlying conditions. Not being fully vaccinated against COVID-19 and/or without a booster remained as a significant modifiable risk factor in the period when Omicron (BA.1) was the dominant strain. These findings may inform medical decisions and help prioritize resources allocations.

## Supporting information

RECORD checklist

## Data Availability

Data in the present study are available upon reasonable request at: https://www.alberta.ca/health-research.aspx

## References

1. Tasker J. Health Canada approves Pfizer’s COVID-19 antiviral treatment: CBC News; 2022 [updated 2022-01-17; cited 2022. Available from: https://www.cbc.ca/news/politics/health-canada-pfizer-therapeutic-1.6317505.

2. Government of Canada. Regulatory Decision Summary - Paxlovid - Health Canada 2022 [cited 2022. Available from: https://covid-vaccine.canada.ca/info/regulatory-decision-summary-detail.html?linkID=RDS00904.

3. Pfizer Inc. Pfizer’s Novel COVID-19 Oral Antiviral Treatment Candidate Reduced Risk of Hospitalization or Death by 89% in Interim Analysis of Phase 2/3 EPIC-HR Study 2021 [updated 2021-11-05; cited 2022. Available from: https://www.pfizer.com/news/press-release/press-release-detail/pfizers-novel-covid-19-oral-antiviral-treatment-candidate.

4. Pfizer Inc. Product Monograph: PAXLOVID. Government of Canada 2022-01-17.

5. Leung K, Jit M, Leung GM, Wu JT. The allocation of COVID-19 vaccines and antivirals against emerging SARS-CoV-2 variants of concern in East Asia and Pacific region: A modelling study. The Lancet Regional Health-Western Pacific. 2022;21:100389.

6. Diep AN, Gilbert A, Saegerman C, Gangolf M, D’Orio V, Ghuysen A, et al. Development and validation of a predictive model to determine the level of care in patients confirmed with COVID-19. Infectious Diseases. 2021;53(8):590–9.

7. Gershengorn HB, Patel S, Shukla B, Warde PR, Bhatia M, Parekh D, et al. Association of race and ethnicity with COVID-19 test positivity and hospitalization is mediated by socioeconomic factors. Annals of the American Thoracic Society. 2021;18(8):1326–34.

8. Sperger J, Shah KS, Lu M, Zhang X, Ungaro RC, Brenner EJ, et al. Development and validation of multivariable prediction models for adverse COVID-19 outcomes in patients with IBD. BMJ Open. 2021;11(11):e049740.

9. Vila-Corcoles A, Satue-Gracia E, Vila-Rovira A, de Diego-Cabanes C, Forcadell-Peris MJ, Ochoa-Gondar O. Development of a predictive prognostic rule for early assessment of COVID-19 patients in primary care settings. Atención Primaria. 2021;53(9):102118.

10. Webb BJ, Levin NM, Grisel N, Brown SM, Peltan ID, Spivak ES, et al. Simple scoring tool to estimate risk of hospitalization and mortality in ambulatory and emergency department patients with COVID-19. PLOS ONE. 2022;17(3):e0261508.

11. Yamada G, Hayakawa K, Matsunaga N, Terada M, Suzuki S, Asai Y, et al. Predicting respiratory failure for COVID-19 patients in Japan: a simple clinical score for evaluating the need for hospitalisation. Epidemiology & Infection. 2021;149.

12. Jehi L, Ji X, Milinovich A, Erzurum S, Merlino A, Gordon S, et al. Development and validation of a model for individualized prediction of hospitalization risk in 4,536 patients with COVID-19. PLOS ONE. 2020;15(8):e0237419.

13. Knight SR, Ho A, Pius R, Buchan I, Carson G, Drake TM, et al. Risk stratification of patients admitted to hospital with COVID-19 using the ISARIC WHO Clinical Characterisation Protocol: development and validation of the 4C Mortality Score. BMJ. 2020;370.

14. Wargny M, Potier L, Gourdy P, Pichelin M, Amadou C, Benhamou P-Y, et al. Predictors of hospital discharge and mortality in patients with diabetes and COVID-19: Updated results from the nationwide CORONADO study. Diabetologia. 2021;64(4):778–94.

15. Kalafat E, Prasad S, Birol P, Tekin AB, Kunt A, Di Fabrizio C, et al. An internally validated prediction model for critical COVID-19 infection and intensive care unit admission in symptomatic pregnant women. American Journal of Obstetrics and Gynecology. 2022;226(3):403. e1-. e13.

16. Tang X, Gelband H, Nagelkerke N, Bogoch II, Brown P, Morawski E, et al. COVID-19 vaccination intention during early vaccine rollout in Canada: A nationwide online survey. The Lancet Regional Health-Americas. 2021;2:100055.

17. Staff A. A timeline of COVID-19 vaccine developments in 2021. American Journal of Managed Care https://www ajmc com/view/a-timel ine-of-covid-19-vacci ne-deve l opmen ts-in-2021 Published June. 2021;3.

18. Baraniuk C. COVID-19: How the UK vaccine rollout delivered success, so far. BMJ. 2021;372.

19. Sheikh A, McMenamin J, Taylor B, Robertson C. SARS-CoV-2 Delta VOC in Scotland: demographics, risk of hospital admission, and vaccine effectiveness. The Lancet. 2021;397(10293):2461–2.

20. Sheikh A, Robertson C, Taylor B. BNT162b2 and ChAdOx1 nCoV-19 vaccine effectiveness against death from the delta variant. New England Journal of Medicine. 2021;385(23):2195–7.

21. Vasileiou E, Simpson CR, Robertson C, Shi T, Kerr S, Agrawal U, et al. Effectiveness of first dose of COVID-19 vaccines against hospital admissions in Scotland: National prospective cohort study of 5.4 million people. 2021.

22. Paredes MI, Lunn SM, Famulare M, Frisbie LA, Painter I, Burstein R, et al. Associations between SARS-CoV-2 variants and risk of COVID-19 hospitalization among confirmed cases in Washington State: A retrospective cohort study. medRxiv. 2021.

23. Ontario Agency for Health Protection and Promotion (Public Health Ontario). COVID-19 variant of concern Omicron (B.1.1.529): Risk assessment. Toronto, ON: Queen’s Printer for Ontario 2022-02-11.

24. Auvigne V, Vaux S, Le Strat Y, Schaeffer J, Fournier L, Montagnat C, et al. Serious hospital events following symptomatic infection with Sars-CoV-2 Omicron and Delta variants: An exposed-unexposed cohort study in December 2021 from the COVID-19 surveillance databases in France. medRxiv. 2022.

25. Peralta-Santos A, Leite PP, Casaca P, Fernandes E, Rodrigues EF, Moreno J, et al. Omicron (BA. 1) SARS-CoV-2 variant is associated with reduced risk of hospitalization and length of stay compared with Delta (B. 1.617. 2). medRxiv. 2022.

26. Cunningham CT, Cai P, Topps D, Svenson LW, Jetté N, Quan H. Mining rich health data from Canadian physician claims: features and face validity. BMC Res Notes. 2014;7(1):1–8.

27. Tran DT, Palfrey D, Lo T, Welsh R. Outcome and Cost of Optimal Control of Dyslipidemia in Adults With High Risk for Cardiovascular Disease. Can J Cardiol. 2021;37(1):66–76.

28. Marshall DA, Liu X, Barnabe C, Yee K, Faris PD, Barber C, et al. Existing comorbidities in people with osteoarthritis: a retrospective analysis of a population-based cohort in Alberta, Canada. BMJ Open. 2019;9(11):e033334.

29. McAlister FA, Wiebe N, Hemmelgarn BR. Time in therapeutic range and stability over time for warfarin users in clinical practice: A retrospective cohort study using linked routinely collected health data in Alberta, Canada. BMJ Open. 2018;8(1):e016980.

30. Fatoye F, Gebrye T, Svenson LW. Real-world incidence and prevalence of systemic lupus erythematosus in Alberta, Canada. Rheumatol Int. 2018;38(9):1721–6.

31. Chen G, Faris P, Hemmelgarn B, Walker RL, Quan H. Measuring agreement of administrative data with chart data using prevalence unadjusted and adjusted kappa. BMC Med Res Methodol. 2009;9(1):1–8.

32. Carrillo-Vega MF, Salinas-Escudero G, García-Peña C, Gutiérrez-Robledo LM, Parra-Rodríguez L. Early estimation of the risk factors for hospitalization and mortality by COVID-19 in Mexico. PLOS ONE. 2020;15(9):e0238905.

33. Cottini M, Lombardi C, Berti A, Gregis M, Gregis G, Bello L, et al., editors. Obesity is a major risk factor for hospitalization in community-managed COVID-19 pneumonia. Mayo Clinic Proceedings; 2021: Elsevier.

34. Ko JY, Danielson ML, Town M, Derado G, Greenlund KJ, Kirley PD, et al. Risk factors for COVID-19-associated hospitalization: COVID-19-associated hospitalization surveillance network and behavioral risk factor surveillance system. medRxiv. 2020.

35. Kompaniyets L, Goodman AB, Belay B, Freedman DS, Sucosky MS, Lange SJ, et al. Body mass index and risk for COVID-19–related hospitalization, intensive care unit admission, invasive mechanical ventilation, and death—United States, March–December 2020. Morbidity and Mortality Weekly Report. 2021;70(10):355.

36. Soares RdCM, Mattos LR, Raposo LM. Risk factors for hospitalization and mortality due to COVID-19 in Espírito Santo State, Brazil. American Journal of Tropical Medicine and Hygiene. 2020;103(3):1184.

37. Vahey GM, McDonald E, Marshall K, Martin SW, Chun H, Herlihy R, et al. Risk factors for hospitalization among persons with COVID-19—Colorado. PLOS ONE. 2021;16(9):e0256917.

38. Thompson MG, Stenehjem E, Grannis S, Ball SW, Naleway AL, Ong TC, et al. Effectiveness of COVID-19 vaccines in ambulatory and inpatient care settings. New England Journal of Medicine. 2021.

39. Thompson MG, Stenehjem E, Grannis S, Ball SW, Naleway AL, Ong TC, et al. Effectiveness of COVID-19 vaccines in ambulatory and inpatient care settings: Protocol. 2021.

40. Guan W-j, Liang W-h, Zhao Y, Liang H-r, Chen Z-s, Li Y-m, et al. Comorbidity and its impact on 1590 patients with COVID-19 in China: A nationwide analysis. European Respiratory Journal. 2020;55(5).

41. Levin EG, Lustig Y, Cohen C, Fluss R, Indenbaum V, Amit S, et al. Waning immune humoral response to BNT162b2 COVID-19 vaccine over 6 months. New England Journal of Medicine. 2021;385(24):e84.

42. Lam C, Calvert J, Siefkas A, Barnes G, Pellegrini E, Green-Saxena A, et al. Personalized stratification of hospitalization risk amidst COVID-19: A machine learning approach. Health policy and technology. 2021;10(3):100554.

43. Alberta Health Services. Assessment & Testing: COVID-19 [2022/1/10]. Available from: https://www.albertahealthservices.ca/topics/Page17058.aspx.

